# “Effectiveness of whole virus COVID-19 vaccine at protecting health care personnel against SARS-CoV-2 infections in Lima, Peru”

**DOI:** 10.1101/2022.03.16.22271100

**Authors:** Carmen S Arriola, Giselle Soto, Matthew Westercamp, Susan Bollinger, Angelica Espinoza, Max Grogl, Alejandro Llanos-Cuentas, Eduardo Matos, Candice Romero, Maria Silva, Rachel Smith, Natalie Olson, Michael Prouty, Eduardo Azziz-Baumgartner, Fernanda C. Lessa

## Abstract

In February 2021, Peru launched a vaccination campaign among healthcare personnel using BBIBP-CorV inactivated whole virus (BBIBP-CorV) COVID-19 vaccine. Two doses of BBIBP-CorV vaccine are recommended, 21 days apart. Data on BBIBP-CorV vaccine effectiveness will inform the use and acceptance of vaccination with BBIBP-CorV vaccine.

We evaluated BBIBP-CorV vaccine effectiveness among an existing multi-year influenza cohort at two hospitals in Lima. We analyzed data on 290 participants followed between February and May 2021. Participants completed a baseline questionnaire and provided weekly self-collected anterior nasal swabs tested for SARS-CoV-2 by rRT-PCR for sixteen weeks. We performed multivariable logistic regression models adjusting for pre-selected characteristics (age, sex, exposure to COVID-19 patients, work in intensive care unit or emergency department, BMI, and exposure time in days). BBIBP-CorV vaccine effectiveness was calculated after the two-week post-vaccination period as (1-Odds Ratio for testing SARS-CoV-2 positive)x100%.

SARS-CoV-2 was detected by rRT-PCR among 25 (9%) participants during follow-up (February-May 2021). Follow-up period ranged 1-11 weeks (median: 2 weeks). Among cohort participants who were fully vaccinated the adjusted vaccine effectiveness against SARS-CoV-2 infection was estimated as 95% (95% CI: 70%, 99%) and 100% (95% CI: 88%, 100%) for those partially vaccinated.

During the study period, vaccination of healthcare personnel with BBIBP-CorV vaccine was effective at reducing SARS-CoV-2 infections in the weeks immediately following vaccination. This information can be used to support vaccination efforts in the region, especially among those who could be concerned about their effectiveness.

## Introduction

Peru is a middle-income country (MIC) disproportionately affected by coronavirus disease (COVID-19) and struggling to protect its essential workforce [1-4]. Despite early lockdowns, curfews, and other public health and social measures implemented to reduce disease spread [5], as of May 22, 2021, Peru had reported 180,764 COVID-19 associated deaths [6], and continues to accrue cases [7]. As in many other MICs, healthcare services have been overwhelmed with patients, have limited personal protective equipment, and have delayed and limited COVID-19 vaccination, leading to unrest and strikes among Peruvian healthcare personnel [8]. COVID-19 vaccination with Sinopharm BBIBP-CorV (Beijing Institute of Biological Products Coronavirus Vaccine) inactivated whole virus vaccine, hereinafter referred to as BBIBP-CorV vaccine, was initiated in Peru on February 9, 2021, with healthcare personnel as a priority group for vaccination. During the study period (February 9 to May 4, 2021), BBIBP-CorV vaccine was the only COVID-19 vaccine available in Peru for healthcare personnel [9, 10]. Two doses of BBIBP-CorV vaccine are recommended, 21 days apart. To date, BBIBP-CorV vaccine effectiveness data against SARS-CoV-2 infection have not been published in the peer-reviewed literature. Evidence on BBIBP-CorV vaccine effectiveness could reduce hesitancy about the vaccine and would support vaccination efforts. We leveraged an existing multi-year influenza cohort of healthcare workers at two Lima hospitals [11] to evaluate BBIBP-CorV vaccine effectiveness at preventing both symptomatic and asymptomatic SARS-CoV-2 infections during the study period.

## Methods

### Study design

From the start of COVID-19 vaccination among healthcare personnel on February 9, 2021 until May 4, 2021, we followed healthcare personnel aged 18 to 65 years who were participating in an ongoing cohort of healthcare workers at two tertiary referral hospitals in Lima [11]. Inclusion criteria for cohort eligibility were: age ≥18 years; work at the facility full-time (≥30 hours per week); have routine, direct, hands-on or face-to-face contact with patients (within 1 meter) as part of a typical work shift; and have worked at the facility for ≥1 year prior to enrollment.

Between November 25, 2020, and January 9, 2021, participants provided written informed consent and completed a baseline questionnaire about their demographics and role in the hospital (self-report exposure to COVID-19 patients (yes/no), work in intensive care unit (ICU) (yes/no), emergency department (ED) (yes/no)). Participants provided serum samples at baseline and at the end of the study period. Each participant was followed for 16 weeks after enrollment. Participants responded to a weekly survey that included questions about COVID-19 exposure and receipt of BBIBP-CorV vaccine (documented by the hospitals). Participants also provided a weekly self-collected anterior nasal swab which was tested for SARS-CoV-2 by real-time reverse transcription polymerase chain reaction (rRT-PCR) at the Naval Medical Research Unit 6 (NAMRU-6) in Lima, following Centers for Disease Control and Prevention (CDC) testing protocols [12]. rRT-PCR testing was performed in pools of five samples; if positive, all five individual samples were tested separately. Serum samples were shipped to the CDC in Atlanta, Georgia, United States of America, for pan-Ig serology testing [13].

### Statistical analysis

We described healthcare personnel demographics, occupational information, baseline serology, COVID-19 vaccine receipt and laboratory detection of SARS-CoV-2. We applied Chi-square or Wilcoxon tests, as appropriate, to assess differences in demographics, occupational information, and baseline serology, stratified by SARS-CoV-2 detection and COVID-19 vaccine receipt.

We estimated vaccine effectiveness using a multivariable logistic regression model adjusted for pre-selected characteristics [i.e., age, sex, exposure to COVID-19 patients, work in ICU, ED, body mass index (BMI), and time of follow-up in days] and presented vaccine effectiveness estimates [(1-Odds Ratio) x100%] with 95% confidence intervals (CI). For these analyses, we excluded those who were seropositive at baseline and those with a positive COVID-19 test before February 9, 2021. We defined full vaccination as the period starting 14 days after receipt of the second dose and partial vaccination as the period starting 14 days after receipt of the first dose. Participants not meeting these criteria were considered unvaccinated. The partial vaccination model only included those who received one dose of the vaccine during the study period. Partially vaccinated individuals were excluded from the full vaccination analysis. COVID-19 vaccine effectiveness was calculated under both full and partial vaccination scenarios. The outcome of interest in the model was SARS-CoV-2 detection; if SARS-CoV-2 was detected in an individual before first vaccination date or before the two-week period after first vaccination, he/she would be considered unvaccinated for the analysis. All analyses were conducted in R (R version 4.1.0).

### Ethics

This study was reviewed and approved by the NAMRU6 IRB as the single IRB (sIRB) for this study^§^.

## Results

### Study sample characteristics, SARS-CoV-2 infections and COVID-19 vaccine receipt

Two hundred and ninety cohort participants were followed between February 9 and May 4, 2021, of which 270 (93.1%) reported receiving at least one dose of COVID-19 vaccine; of these, 80% (216/270) reported being fully vaccinated before the end of follow-up. The follow-up period after vaccination ranged 1-11 weeks after the two-week post-vaccination period; median of 2 weeks. Median age of participants was 45 years (IQR: 38, 52). Three out of four (74%; 215/290) participants were female and 90% (260/290) reported being of mixed race. Only 3% (8/290) reported a chronic medical condition, including asthma, diabetes, high blood pressure, chronic heart disease, autoimmune condition, HIV/AIDS, or other medical condition requiring clinical care for at least 6 months. Of 290 participants, 49% (n=143) were classified as overweight and 22% (n=64) as obese. One third (106/290) of participants’ baseline sera had a reactive result for SARS-CoV-2 pan-Ig antibodies, and SARS-CoV-2 was detected by rRT-PCR among 25 (9%) participants during follow-up. Those seronegative at baseline were more likely to subsequently test positive for SARS-CoV-2 through rRT-PCR than those seropositive at baseline (p-value<0.001; Table 1).

**Table 1.** Demographic, occupational information, baseline serology, COVID-19 vaccine receipt and laboratory detection of SARS-CoV-2 among healthcare personnel cohort participants, Lima-Peru, February 9 to May 4, 2021, N=290

|  | Total<br>N=290 (100%) | SARS-CoV-2<br>positive * | SARS-CoV-2<br>negative | p-value□ | Unvaccinated<br>n=20 (7%) | Partially<br>vaccinated**<br>n=54 (19%) | p-value□ | Fully vaccinated**<br>n=216 (74%) | p-value□ |
| --- | --- | --- | --- | --- | --- | --- | --- | --- | --- |
| Age (years) |  |  |  |  |  |  |  |  |  |
| Median (IQR) | 45 (38,52) | 48 (41, 54) | 45 (38,51) |  | 39 (37,49) | 47 (39, 52) |  | 45 (39,52) |  |
| 18-39 | 85 (29) | 6 (24) | 79 (30) | 0.82 | 10 (50) | 14 (26) | 0.14 | 61 (28) | 0.12 |
| 40-49 | 110 (38) | 10 (40) | 100 (38) |  | 5 (25) | 22 (41) |  | 83 (38) |  |
| 50-65 | 95 (33) | 9 (36) | 86 (32) |  | 5 (25) | 18 (33) |  | 72 (33) |  |
| Gender |  |  |  |  |  |  |  |  |  |
| Male | 75 (26) | 6 (24) | 69 (26) | 1.0 | 1 (5) | 25 (46) | <0.01 | 49 (23) | 0.12 |
| Female | 215 (74) | 10 (76) | 196 (74) |  | 19 (95) | 29 (54) |  | 167 (77) |  |
| Race/Ethnicity |  |  |  |  |  |  |  |  |  |
| Mixed race | 260 (90) | 21 (84) | 239 (90) | 0.45 | 19 (95) | 45 (83) | 0.48 | 196 (91) | 0.89 |
| Indigenous | 19 (7) | 2 (8) | 17 (6) |  | 1 (5) | 3 (6) |  | 15 (7) |  |
| Black | 8 (3) | 1 (4) | 7 (3) |  | 0 (0) | 5 (9) |  | 3 (1) |  |
| White | 3 (1) | 1 (4) | 2 (1) |  | 0 (0) | 1 (2) |  | 2 (1) |  |
| Education |  |  |  |  |  |  |  |  |  |
| High school<br>education only | 37 (13) | 2 (8) | 35 (13) | 0.75 | 0 (0) | 13 (24) | 0.02 | 24 (11) | 0.10 |
| Associate degree<br>from college or<br>equivalent/Bachelor's degree | 233 (80) | 21 (84) | 212 (80) |  | 20 (100) | 38 (70) |  | 175 (81) |  |
| Postgraduate<br>education | 20 (7) | 2 (8) | 18 (7) |  | 0 (0) | 3 (6) |  | 17 (8) |  |
| Comorbidities |  |  |  |  |  |  |  |  |  |
| Any medical<br>condition** | 8 (3) | 2 (8) | 6 (2) | 0.52 | 1 (5) | 2 (4) | 0.74 | 5 (2) | 0.64 |
| BMI |  |  |  |  |  |  |  |  |  |
| Normal (18.5 to <25) | 83 (29) | 9 (36) | 74 (28) |  | 9 (45) | 17 (31) |  | 57 (26) |  |
| Overweight (25 to <30) | 143 (49) | 12 (48) | 131 (49) | 0.61 | 7 (35) | 27 (50) | 0.47 | 109 (50) | 0.20 |
| Obese ($\geq 30$) | 64 (22) | 4 (16) | 60 (23) | | 4 (20) | 10 (19) | | 50 (23) | |
| Smoking (daily/some) | 11 (4) | 1 (4) | 10 (4) | 1.0 | 1 (5) | 3 (6) | 1.0 | 7 (3) | 1.0 |
| Job type |  |  |  |  |  |  |  |  |  |
| Physician | 11 (4) | 1 (4) | 10 (4) |  | 0 (0) | 0 (0) |  | 11 (5) |  |
| Nurse | 63 (22) | 2 (8) | 61 (23) |  | 1 (5) | 10 (19) |  | 52 (24) |  |
| Midwife/Dentist | 12 (4) | 1 (4) | 11 (4) |  | 0 (0) | 0 (0) |  | 12 (5) |  |
| Technician/assistant | 135 (47) | 14 (56) | 121 (46) |  | 15 (75) | 17 (31) |  | 103 (48) |  |
| Pharmacist/social worker/nutritionist | 2 (1) | 0 (0) | 2 (1) | 0.62 | 1 (5) | 0 (0) | <0.01 | 1 (0) | 0.08 |
| Physical therapist | 4 (1) | 0 (0) | 4 (2) |  | 0 (0) | 2 (4) |  | 2 (1) |  |
| Admin/security/maintenance/Transporter | 49 (17) | 7 (28) | 42 (15) |  | 1 (5) | 20 (37) |  | 28 (12) |  |
| Other | 14 (5) | 0 (0) | 14 (5) |  | 2 (10) | 5 (9) |  | 7 (3) |  |
| Exposed to COVID-19 Patients in healthcare setting | 249 (86) | 18 (72) | 231 (87) | 0.59 | 17 (85) | 45 (83) | 1.0 | 187 (87) | 1.0 |
| Work in ICU | 27 (9) | 4 (16) | 23 (9) | 0.40 | 0 (0) | 8 (15) | 0.16 | 19 (9) | 0.34 |
| Work in Emergency Department | 101 (35) | 11 (44) | 90 (34) | 0.43 | 9 (45) | 20 (37) | 0.72 | 72 (33) | 0.42 |
| Number of work hours at site per week (median, IQR) | 36 (36,36) | 36 (36,40) | 36 (36,36) | 0.93 | 36 (36,39) | 36 (36,48) | 0.23 | 36 (36,36) | 0.40 |
| Number of patient-provider face-to-face hours per week (median, IQR) | 30 (24,36) | 30 (20,30) | 30 (25,36) | 0.04 | 33 (30,37) | 30 (25,36) | 0.31 | 30 (24,36) | 0.11 |
| Reactive SARS-CoV-2 serology at baseline | 106 (37) | 0 (0) | 106 (40) | <0.001 | 5 (25) | 15 (28) | 1.0 | 86 (40) | 0.28 |

Notes:
\* At least once by weekly testing during follow-up period
\*\*Vaccination of healthcare workers started in Lima on February 9, 2021 and was assessed by interview on a weekly basis. Partially vaccinated refers to those who received 1 dose of whole virus COVID-19 vaccine during the study period; fully vaccinated refers to those who received 2 doses of whole virus COVID-19 vaccine during the study period
\*\*\*Asthma, diabetes, high blood pressure, chronic heart disease, autoimmune condition, HIV/AIDS, another medical condition requiring clinical care for at least 6 months
□ Chi-square test for categorical and Wilcoxon signed rank test for continuous variables. Partially and fully vaccinated groups were compared to unvaccinated individuals, separately

### COVID-19 vaccine effectiveness

After excluding those seropositive at baseline and those with a positive COVID-19 test before February 9, 2021, and adjusting for age, sex, exposure to COVID-19 patients, work in ICU, work in ED, BMI, and time of follow-up in days, COVID-19 vaccine effectiveness against SARS-CoV-2 infection (either symptomatic or asymptomatic) was estimated as 97% (95% CI: 88%, 99%) for those who received at least one dose of the vaccine; 100% (95% CI: 88%, 100%) for those partially vaccinated, and 95% (95% CI: 70%, 99%) for those fully vaccinated (Table 2).

**Table 2.** COVID-19 vaccine effectiveness (VE) by dose received among healthcare personnel cohort participants, Lima-Peru, February 9 to May 4, 2021

|  | COVID-19 Cases |  | Controls |  | Unadjusted VE<br>% (95% CI) | Adjusted VE*<br>% (95% CI) |
| --- | --- | --- | --- | --- | --- | --- |
|  | Vaccinated | Unvaccinated | Vaccinated | Unvaccinated |  |  |
| *Received at least one dose of vaccine* | 10 | 9 | 138 | 6 | 95% (84%, 99%) | 97% (88%, 99%) |
| *Fully vaccinated* | 5 | 9 | 36 | 6 | 91% (63%, 98%) | 95% (70%, 99%) |
| *Partially vaccinated* | 5 | 9 | 25 | 6 | 87% (45%, 97%) | 100% (88%, 100%) |

\*Adjusted for age, sex, exposure to COVID-19 patients, work in ICU, work in ER, BMI, and time of follow-up.
Note1: Those with reactive SARS-CoV-2 serology at baseline have been excluded (n=106).
Note2: Those with a positive COVID-19 test before February 9, 2021 have been excluded (n=17).
Note3: Those who had tested positive before vaccination date or before the 2-week period after vaccination were considered unvaccinated for the model.
Note 4: We defined full vaccination as the period starting 14 days after receipt of the second dose and partial vaccination as the period starting 14 days after receipt of the first dose. Participants not meeting these criteria were considered unvaccinated. The partial vaccination model only included those who received one dose of the vaccine during the study period.

## Discussion

Among those vaccinated in this cohort, we estimate BBIBP-CorV vaccine was ≥90% effective in preventing infection with SARS-CoV-2 in the weeks immediately following vaccination. To our knowledge, this is the first peer-reviewed data of BBIBP-CorV vaccine effectiveness against SARS-CoV-2 infection. Furthermore, our findings indicate that during the study period (February to May 2021), one in ten study participants in two tertiary hospitals in Lima were infected with laboratory-confirmed SARS-CoV-2.

Healthcare personnel are at increased risk of becoming infected with SARS-CoV-2 [14]. Our findings show continued detection of SARS-CoV-2 infection during the study period. In Peru, it is estimated that >600 physicians and nurses have died of COVID-19 through June 2021 [1]. Protecting the healthcare workforce is a global priority to ensure delivery of healthcare to the population. The World Health Organization (WHO) Strategic Advisory Group of Experts on Immunization (SAGE) roadmap for prioritizing use of COVID-19 vaccines in the context of limited supplies includes healthcare personnel as one of the highest priority groups for vaccination [15]. The government of Peru initiated their COVID-19 vaccination on February 9, 2021, with healthcare personnel being the initial targeted group to receive the vaccine [16].

Our study indicates the BBIBP-CorV vaccine is effective against SARS-CoV-2 infection in the immediate period after vaccination. Our findings are compatible with those reported by WHO where BBIBP-CorV vaccine efficacy was estimated as 78.9% (95% CI: 65.8%-87%) against COVID-19 disease in an unpublished clinical trial, with a follow-up time of two months [17]. Furthermore, our findings are consistent with interim estimates published by WHO, which estimated that vaccine effectiveness against rt-PCR confirmed cases among adults 18 years of age and older in Bahrain was 90% (95% CI: 88%, 91%) [18].

In our study, we suspect that B.1.1.1 was the dominant circulating variant in early 2021, as it was detected in 43% of the samples that were sequenced (n=23). However, SARS-CoV-2 variant P.1 was identified for the first time in Peru in January 2021 [19] and P.1 was identified in one out of 19 samples that were collected in January and February 2021 (data not shown); this strain emerged in Brazil in mid-November and rapidly spread in the state of Amazonas in early 2021, causing several hospitalizations and deaths [20, 21]. WHO included P.1 as one of four variants of concern in January 2021 because of its increased transmissibility and virulence [22]. It is important to maintain data collection over time to assess vaccine effectiveness under real-life circumstances while new variants circulate.

Due to high COVID-19 morbidity and mortality, BBIBP-CorV vaccine was rolled out in Peru and numerous other countries despite not having robust effectiveness data [23]. Though long-term effectiveness data are still needed, the results from our study support BBIBP-CorV’s continued use, at least in the absence of available vaccines with proven long-term effectiveness. Data from this study can be used to support vaccination in the region; presenting vaccine effectiveness data can improve vaccine uptake [24]. Unlike some other COVID-19 vaccines, BBIBP-CorV has the advantage that it does not require complicated cold chain logistics, such as ultralow freezer conditions; instead, it can be used within the existing cold chain infrastructure of national immunization programs [25].

This study had several noteworthy strengths. This prospective cohort study was rapidly implemented by leveraging an ongoing prospective cohort established to evaluate influenza vaccine effectiveness among healthcare personnel with weekly swabbing and testing for SARS-CoV-2, regardless of symptoms. This frequency and breadth of sampling among our cohort allowed for greater detection of infection than in passive surveillance systems. Participation rate in this COVID-19 study was high (85%) and remained high throughout the 16-week follow-up period (>96% of participants submitted specimens in at least 13 of the 16 weeks of follow-up). SARS-CoV-2 infection was confirmed through rRT-PCR in NAMRU-6’s high proficiency laboratory, following CDC’s SARS-CoV-2 diagnostic protocol, and did not rely on point of care testing with less sensitive assays.

This study had at least four limitations. The high vaccine effectiveness we observed may be related to the short follow-up period after vaccination (range: 1-11 weeks after the two-week post-vaccination period; median of 2 weeks); a longer follow-up period is necessary to fully evaluate the long-term effectiveness of the vaccine among this study population. In addition, the study was not sufficiently powered to stratify VE estimates by variant or symptomatic vs asymptomatic infection. Moreover, because of the limited availability of laboratory staff and high volume of weekly respiratory specimens, we implemented a pooling strategy for SARS-CoV-2 testing which may have decreased sensitivity to detect those with low viral shedding. Last, our study could not distinguish nasal carriage of the virus from lower respiratory tract infection.

In summary, one in ten healthcare personnel in our study in Peru tested positive for SARS-CoV-2 during February to May 2021. Vaccination of healthcare personnel with BBIBP-CorV vaccine was effective at reducing SARS-CoV-2 infections in the weeks immediately following vaccination. The data presented in this study supports the Peruvian Government’s ongoing COVID-19 vaccination efforts for reducing SARS-CoV-2 infections, especially among this critical workforce of healthcare professionals.

## Data Availability

All data produced in the present study are available upon reasonable request to the authors and upon approval from involved institutions.

## Disclaimer

The findings and conclusions in this report are those of the authors and do not necessarily represent the views of the US Centers for Disease Control and Prevention, the Department of the Navy, Department of Defense, nor the US Government.

## Acknowledgments

We thank the participants who enrolled in the study; Nia Mims for the establishment of the CDC-DOD Interagency Agreement; the International Reagent Resource Team at CDC, Influenza Division; the CDC COVID-19 response teams: International Task Force, Laboratory Task Force. We also thank Miriam Gonzalez and Sayda La Rosa for supervision and work with field workers collecting data and samples; Fiorela Alvarez and Rocio Mananita for their clinical support during surveillance activities; NAMRU-6 Laboratory Team, LCDR Paul Graf, LCDR Stephen Lizewski, LT Eugenio Abente, Roger Castillo, Anilu Tecco, for their extraordinary support and hard work processing samples; Vicky Arnao for administrative support.

## Financial Disclosure

The study was funded through CDC Interagency Agreement 19FED1916949IPD.

The study protocol was approved by the U.S. Naval Medical Research Unit 6 (NAMRU-6) Institutional Review Board in compliance with all applicable federal regulations governing the protection of human subjects.

## Copyright Statement

I am a federal employee of the United States government. This work was prepared as part of my official duties. Title 17 U.S.C. 105 provides that ‘copyright protection under this title is not available for any work of the United States Government.’ Title 17 U.S.C. 101 defines a U.S. Government work as work prepared by a military service member or employee of the U.S. Government as part of that person’s official duties.

## Footnotes

§ See 45 C.F.R. part 46.114; 21 C.F.R. part 56.114

